# Ten-Second Cold Water Stress Test Differentiates Parkinson’s Disease From Multiple System Atrophy: A Pilot Study

**DOI:** 10.1101/2024.12.20.24319416

**Authors:** Makoto Takahashi, Wataru Hagiwara, Sakiko Itaya, Keisuke Abe, Tetsuya Maeda, Akira Inaba, Satoshi Orimo

**Affiliations:** Department of Neurology, Kanto Central Hospital, Tokyo, Japan; Division of Neurology and Gerontology, Department of Internal Medicine, School of Medicine, Iwate Medical University, Iwate, Japan; Kamiyoga Setagaya Street Clinic, Tokyo, Japan

**Keywords:** Parkinson’s disease, multiple system atrophy, cold stress test, thermography, surface temperature

## Abstract

**Background:** Patients with Parkinson’s disease (PD) often have cold hands and experience frostbite. The diagnostic criteria for multiple system atrophy (MSA) also describe cold and discolored hands, but in our clinical experience we have noticed that the hands are relatively warm. These symptoms are thought to be caused by autonomic dysfunction; however, the detailed mechanisms and differences in cold hands between MSA and PD remain unclear.

**Objectives:** To identify an appropriate cold stimulation test to differentiate patients with PD and MSA using finger surface temperature (FST).

**Methods:** We included 27 and seven patients diagnosed with PD and MSA, respectively, at least 5 years after disease onset. After 15 minutes in a room with constant temperature and humidity, the patient’s hand was placed in cold water at 4°C for 10 seconds as the cold water stress test (10sec-CWST). FST was captured using a thermal imaging camera every minute for 15 minutes, and the recovery of FST was analyzed. The association between the clinical characteristics of each patient and the degree of FST recovery was examined.

**Results:** All patients completed the 10sec-CWST without adverse events. Patients with PD showed a significantly slower recovery of FST after 7 minutes than that of those with MSA, with a maximum difference at 11 minutes (PD: 8.1±0.6°C; MSA: 10.5±0.3°C; p<0.01). FST recovery at 11 minutes was negatively correlated with the degree of resting hand tremor (r=-0.585, p<0.01).

**Conclusions:** FST after 10sec-CWST may be safe and efficient test to differentiate PD and MSA.

## Introduction

Parkinson’s disease (PD), the second most common neurodegenerative disorder, is characterized by the loss of dopaminergic neurons in the substantia nigra, causing extrapyramidal symptoms.^1,2^ Furthermore, the autonomic nervous system (ANS) is disturbed in PD, and constipation, an autonomic symptom, is observed in early onset or before motor symptoms develop.^1^ Multiple system atrophy (MSA) is a neurodegenerative disorder characterized by central autonomic dysfunction, extrapyramidal symptoms, or cerebellar ataxia.^3^ The parkinsonian variant of MSA (MSA-P), in which parkinsonian symptoms predominate, is clinically similar to PD.^4^ Differentiation between PD and MSA is based on the presence of olfactory disturbance^5^ and cerebellar ataxia,^3^ timing of the onset of falls, speed of progression, and abnormal findings on head magnetic resonance imaging; however, it is often difficult to differentiate between the two diseases, especially in the early stages of onset.

Patients with PD and MSA show autonomic symptoms even in the early phase; however, the degree and pattern of autonomic failure differ clinically and pathologically between these two diseases, and these differences provide clues for differentiation. Pathologically, the autonomic disturbance in PD is predominantly present in the peripheral ANS, whereas that in MSA is predominantly present in the central ANS.^6^ Cardiac metaiodobenzylguanidine (MIBG) scintigraphy, which indicates the degree of peripheral cardiac sympathetic denervation,^7^ shows a strong decrease in accumulation in PD, whereas almost no decrease in accumulation in MSA is shown until the advanced stage.^8,9^ Clinically, patients with MSA show more severe orthostatic hypotension than that of those with PD,^10^ and in terms of urinary disturbance, PD is primarily associated with urinary frequency, whereas MSA is often associated with urinary retention.^3,11^

A large number of patients with PD show cold hands and feet, which suggests autonomic dysfunction,^12^ and some patients also show chilblains in winter. Furthermore, the diagnostic criteria for patients with MSA include “cold, discolored hands and feet” as a supportive non-motor feature.^3^ However, we noticed that patients with MSA have relatively warm hands in our daily practice experience, These symptoms are thought to be caused by autonomic disturbances; however, the precise mechanisms and differences in cold hands between PD and MSA are unclear. Several studies have focused on differentiating PD and MSA by hand temperature, with or without cold stimulation^13,14^; however, no consensus has been reached to date.

Prior to this study, we conducted a pilot study in which healthy participants cooled their hands with an ice pack for 2 minutes. We found that it caused severe pain and that the cooling area was uneven (data not shown). Therefore, the purpose of the current study was to find a simple, safe, and reproducible cold stimulation test to differentiate patients with PD and MSA, and to examine whether there are differences in finger surface temperature (FST) between the two diseases after the cold stress test.

## Methods

### Participants

We included consecutive patients with a clinical diagnosis of PD or MSA who were admitted to Kanto Central Hospital and consented to the test. The clinical diagnoses of PD and MSA were made according to the Movement Disorder Society clinical diagnostic criteria for PD^15^ and the second consensus statement on the diagnosis of MSA,^16^ respectively. Patients who performed the 10-second cold water stress test noted below but whose diagnosis changed within 5 years of onset were excluded from the analysis. We also excluded patients with comorbidities, such as diabetes mellitus, endocrine disorders, intracranial diseases, chronic heart failure, myocardial infarction, and peripheral neuropathies, or medications, such as hormones and autonomic drugs, that could affect FST. Clinical information, including age, sex, disease duration, intensity of motor symptoms, presence and intensity of various nonmotor symptoms, accumulation of cardiac MIBG scintigraphy, levodopa dosage at enrollment, and levodopa equivalent daily dose (LEDD), was collected from medical records during hospitalization.

### Ten-Second Cold Water Stress Test (10sec-CWST)

Before the examination, the participants spent at least 15 minutes relaxing in the examination room, where the temperature was kept at 25°C and the humidity at 50%. After a resting period, the patient’s hand on the side with the more severe akinesia was immersed in cold water at 4°C for 10 seconds. After wiping off the cold water, both hands were photographed every minute for 15 minutes using a thermal imaging camera (FLIR E5; FLIR Systems, Wilsonville, OR, USA). FST was measured just proximal to the middle fingernail using dedicated software (FLIR tools, RRID:SCR_016330, version 5.13.17214; FLIR Systems).

### Statistical Analysis

Statistical analyses were performed using EZR^17^ (version 1.67; Saitama Medical Center, Jichi Medical University, Saitama, Japan). The Welch’s t-test and Mann–Whitney U-test were used to compare the PD and MSA groups. The chi-square test was used to compare categorical variables. The relationships between continuous variables were analyzed using Pearson’s correlation coefficient, and nonparametric variables were analyzed using Spearman’s rank correlation coefficient. The sensitivity and specificity of the 10sec-CWST in differentiating PD from MSA were examined by creating a receiver operating characteristic curve. Two-sided p-values less than 0.05 were considered statistically significant.

## Results

### Patients’ Background Data

Twenty-seven patients with PD and eight with MSA underwent the 10sec-CWST without serious adverse events. One patient with MSA was excluded from the analysis because of a change in the clinical diagnosis from MSA to progressive supranuclear palsy within 5 years of disease onset.

Patient background data are summarized in Table 1. Of the seven patients with MSA, five had cerebellar variants of MSA and two had MSA-P. Only one patient with PD was aware of winter frostbite. The severity of the resting tremor, odor test score (odor stick identification test for the Japanese [OSIT-J]), degree of accumulation on cardiac MIBG scintigraphy, levodopa dosage, and LEDD were significantly different between patients with PD and MSA. FST on the test side immediately before the 10sec-CWST was 1.9°C lower in the PD group than that in the MSA group (p=0.0475).

**Table 1.** Patients’ background. *UPDRS score of the hand on the examining side. The severity of the resting tremor, olfactory score, degree of metaiodobenzylguanidine uptake, levodopa volume, and levodopa equivalent daily dose are significantly different between Parkinson’s disease and multiple system atrophy. PD, Parkinson’s disease; MSA, multiple system atrophy; RBD, REM sleep behavior disorder; OH, orthostatic hypotension; OSIT-J, odor stick identification test for the Japanese; MIBG, metaiodobenzylguanidine; L-dopa, levodopa; LEDD, levodopa equivalent daily dose; FST, finger surface temperature on the examination side.

|  | unit | PD (N=27) | MSA (N=7)<br>MSA-C/P (5/2) | P |
| --- | --- | --- | --- | --- |
| male : female | N | 10:17 | 02:05 | 1. 000 |
| Age (mean) (range) | year | 69.4 (41-88) | 61.9 (52-78) | 0.088 |
| Disease duration (mean) (range) | year | 6.1 (1-17) | 3 (1-5) | 0.090 |
| Hohen–Yahr stage (mean) (range) | - | 2.8 (1-4) | 3 (2-4) | 0.604 |
| resting tremor* (mean) (range) | - | 0.7 (0-2) | 0.1 (0-1) | 0.039 |
| rigidity* (mean) (range) | - | 1.6 (1-2) | 1.0 (0-2) | 0.057 |
| RBD | N | 9 | 2 | 1.000 |
| OH | N | 6 | 4 | 0.157 |
| constipation | N | 20 | 5 | 1.000 |
| tachyuria | N | 15 | 2 | 0.398 |
| dysuria | N | 0 | 4 | 0.157 |
| dyshidrosis | N | 8 | 1 | 0.644 |
| chilblains | N | 1 | 0 | 1.000 |
| OSIT-J (mean) (range) | - | 5.5 (1-11) | 10.5 (9-11) | 0.001 |
| MIBG early (mean) (range) | H/M | 1.92 (1.23-3.28) | 3.22 (3.07-3.52) | 0.009 |
| MIBG delay (mean) (range) | H/M | 1.63 (1.02-2.85) | 2.76 (2.63-2.93) | 0.009 |
| L-dopa (mean) (range) | mg/day | 321.3 (0-800) | 78.57 (0-450) | 0.029 |
| LEDD (mean) (range) | mg/day | 472.35 (0-1440) | 90 (0-450) | 0.018 |
| FST (mean) (range) | °C | 32.9 (26.4-36.7) | 34.8(34.4-35.4) | 0.048 |

\*UPDRS score of the hand on the examining side.

### Change in FST After 10sec-CWST

The 10sec-CWST cooled hands to an average temperature of 10.4°C in patients with PD and MSA. A few patients complained of mild chills during the test. There was a statistical difference between FST at 1-3 minutes after the 10sec-CWST on the examined side and that on the unexamined side at 1-2 and 9-12 minutes (Supplemental Figures 1A and B). When comparing FST recovery temperatures, patients with PD showed a significantly slower recovery of FST than that of patients with MSA from 7-15 minutes after the 10sec-CWST, with a maximum difference at 11 minutes (PD: 8.1±0.6°C; MSA: 10.5±0.3°C, p<0.01) (mean ± standard error) (Figures 1 and 2). When the cut-off line of the recovery FST at 11 minutes was set 9.6°C, the two groups could be differentiated with a sensitivity of 70.3% and specificity of 85.7%, with an area under the curve of 0.786 (Figure 3).

**Figure 1.**
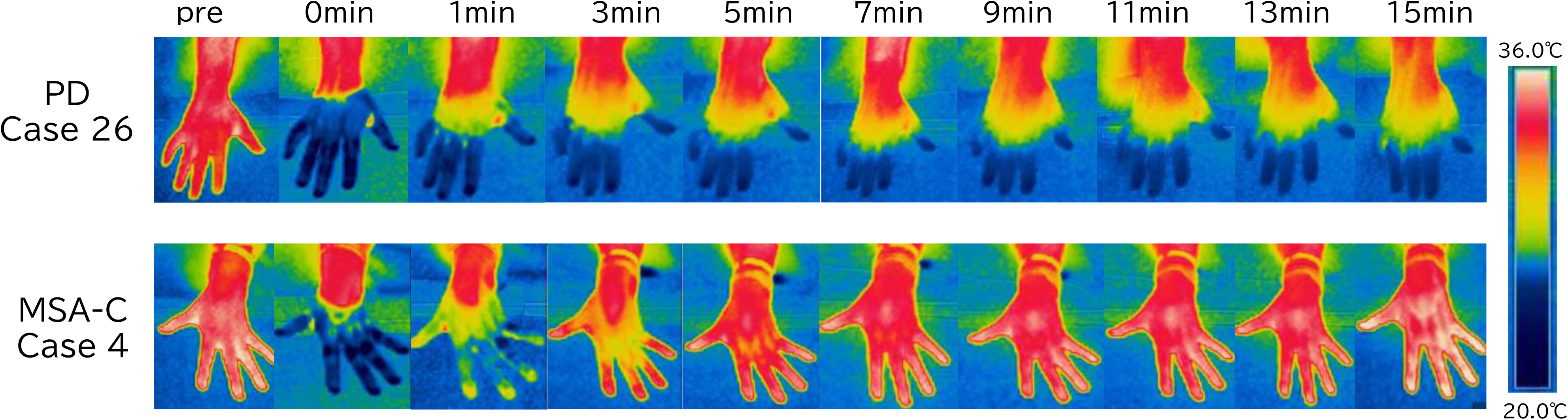
Typical images of finger surface temperature (FST) changes over time in a patient with Parkinson’s disease (PD) and one with multiple system atrophy (MSA). FST in PD (Case 26) rarely recovers, while it shows rapid improvement in MSA (case 4). CWST, cold water stress test; MSA-C, multiple system atrophy – cerebellar subtype; PD, Parkinson’s disease

**Figure 2.**
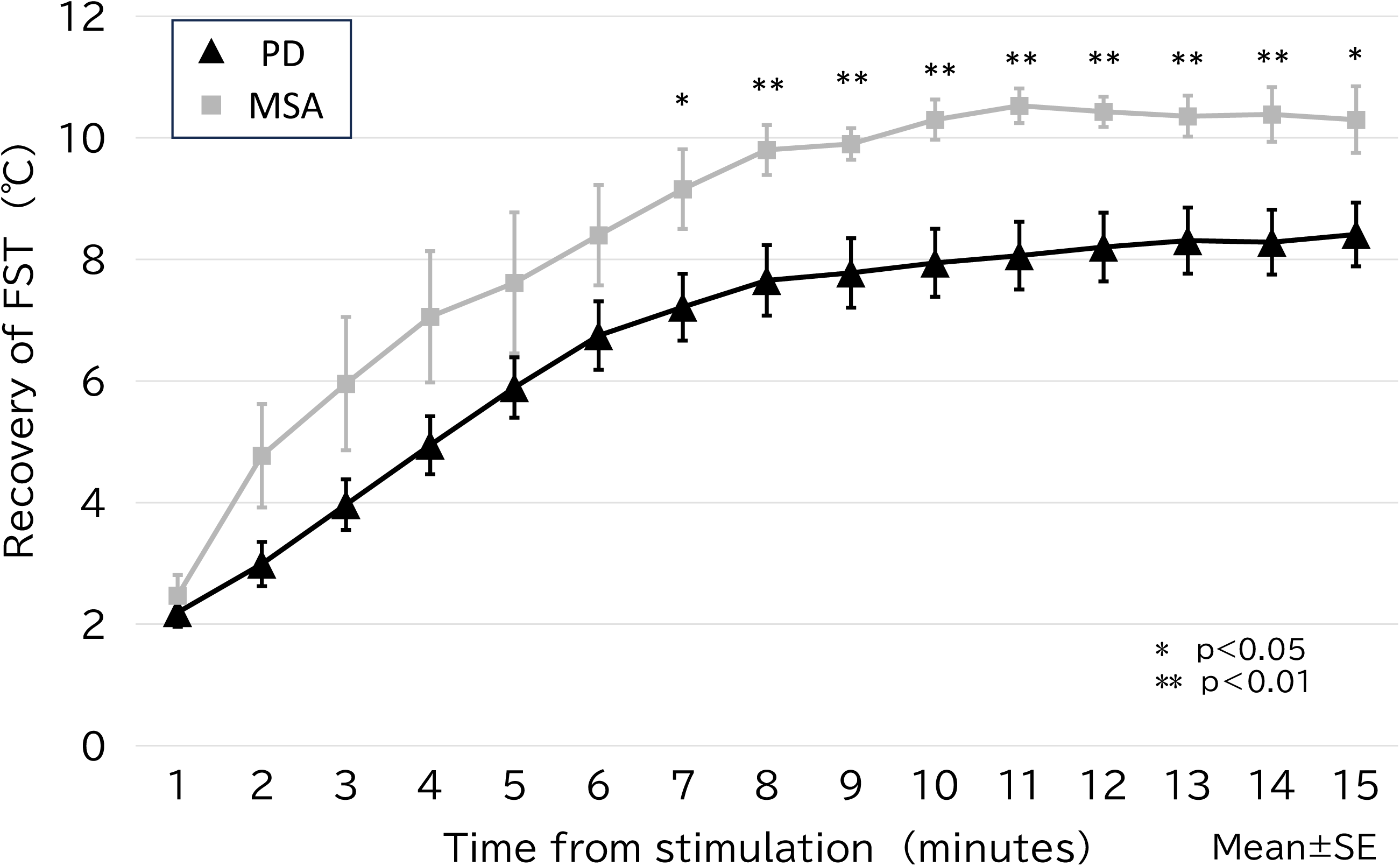
The change in FST over time for the PD and MSA groups. The PD group shows slower FST recovery than that of the MSA group, with a statistically significant difference after 7 minutes of stimulation (maximal difference after 11 minutes). FST, finger surface temperature; MSA, multiple system atrophy; PD, Parkinson’s disease; SE, standard error

**Figure 3.**
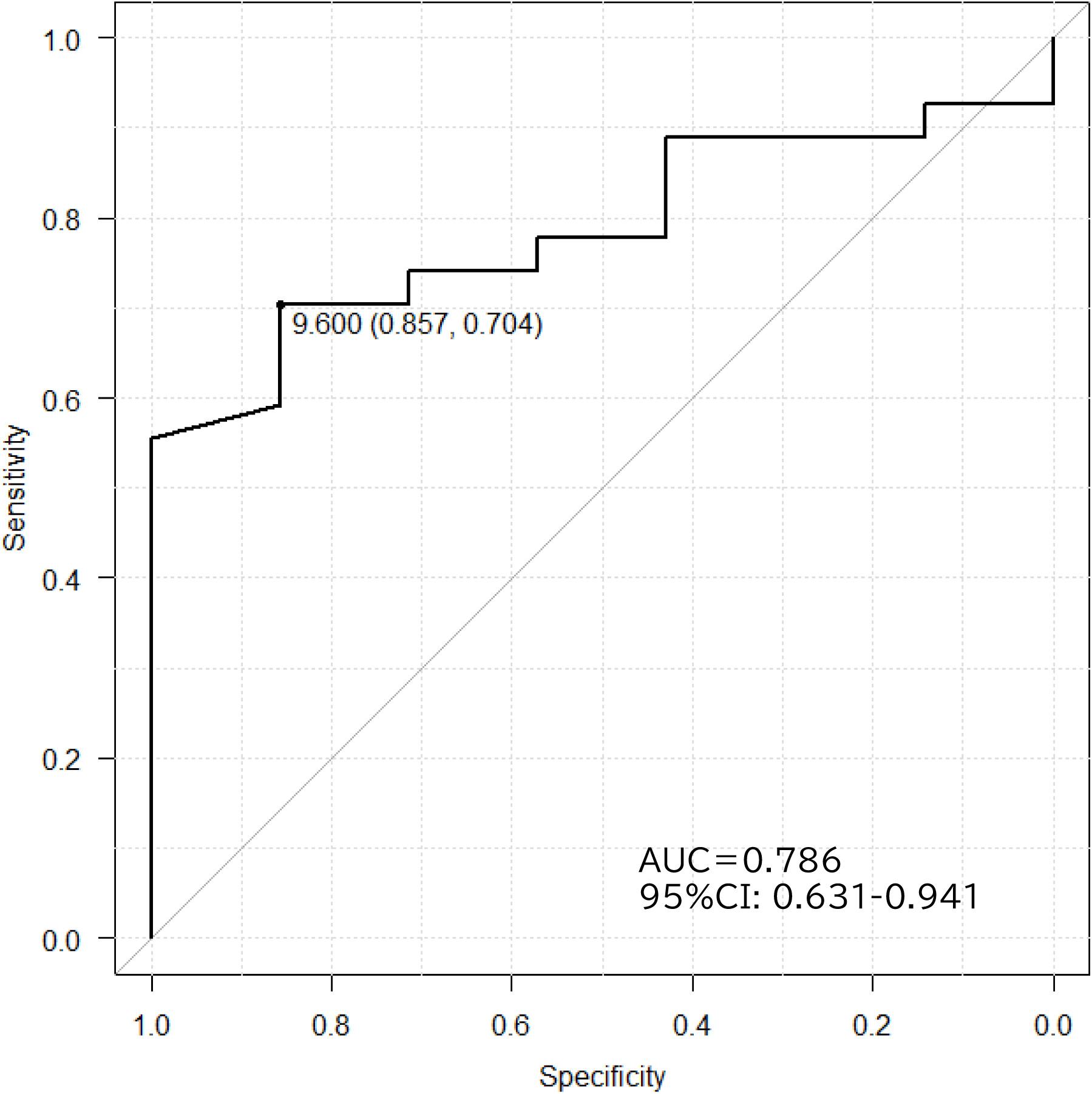
Receiver operating characteristic curves in PD and MSA differentiated by FST recovery after 11 minutes of stimulation. A cutoff value of 9.6°C gives a sensitivity of 70.6%, specificity of 85.7%, and maximum area under the curve of 0.786. AUC, area under the curve; CI, confidence interval

### Factors Related to the Change in FST

In the analysis of all patients, there was a significant correlation between the degree of FST recovery at 11 minutes, degree of resting tremor (r=-0.585, p<0.01), and OSIT-J score (r=0.464, p<0.01) (Figure 4). When the analysis was restricted to the PD group, only the degree of resting tremor showed a statistically significant negative correlation (r=-0.474, p=0.012).

**Figure 4.**
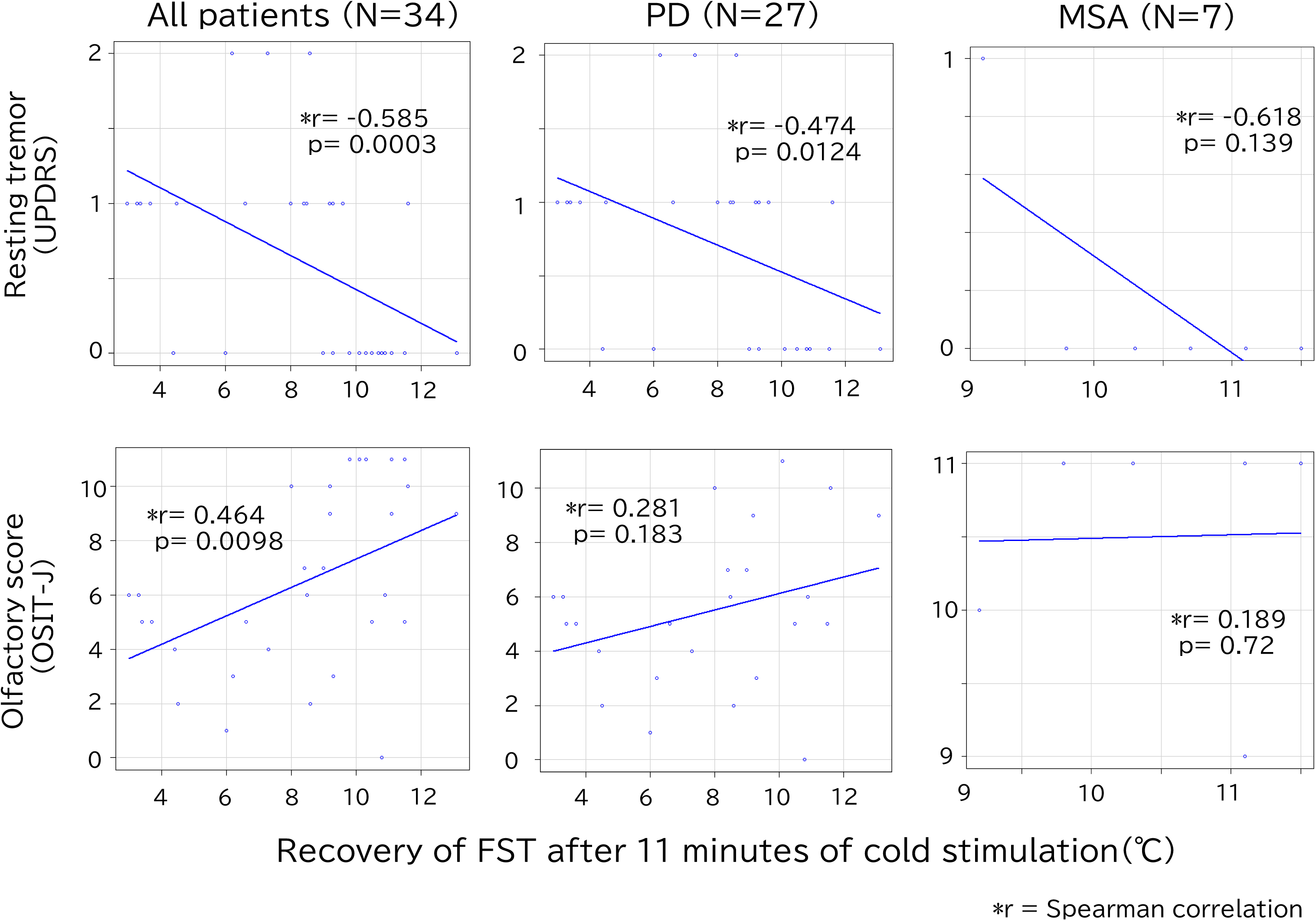
Correlation between recovery of FST after 11 minutes of cold stimulation and resting tremor/olfactory score. In all patients, there is a significant correlation between FST and resting tremor/olfaction scores, but only resting tremor is statistically significant in the PD group. FST, finger surface temperature; MSA, multiple system atrophy; OSIT-J, odor stick identification test for the Japanese; PD, Parkinson’s disease; UPDRS, Unified Parkinson’s Disease Rating Scale

## Discussion

Our study had three important findings: 1) the 10sec-CWST resulted in a sufficiently cool hand uniformly and safely for patients with PD and MSA; 2) although differences were also observed in the pre-stimulus FST between the two groups, a more obvious differentiation between PD and MSA could be made by the degree of change in FST after the 10sec-CWST; and 3) the degree of resting tremor correlated with the degree of change in FST.

The first important finding was that the 10sec-CWST was performed safely and cooled the hands evenly and sufficiently, with no adverse events or dropouts in patients with PD and MSA. The commonly performed cold stress test using icepacks or cold water for more than 1 minute is painful and carries the risk of hyperventilation and dropout.^14,18^ Furthermore, in cold stress tests using icepacks, the contact area between the coolant and skin is non-uniform, resulting in different degrees of cooling in different areas. Therefore, we conducted a cold stress test with cold water to achieve uniform cooling and investigated the minimum time required for a sufficient cooling effect. In the CWST conducted on healthy participants as a pilot test, few were able to tolerate the test for more than 30 seconds; a cooling of approximately 10°C was observed after 10 seconds of CWST, which was considered sufficient for the study. The current study on patients with PD and MSA also showed an average cooling of 10.4°C with the 10sec-CWST, and all patients were able to complete the examination without serious adverse effects, although some complained of a slight chill during CWST.

The second and most important finding was that patients with PD showed a significantly slower recovery of FST than that of those with MSA in the 10sec-CWST and that the 10sec-CWST could distinguish between these two groups based on the degree of FST recovery. The PD group also showed lower FSTs than that of the MSA group before cold stimulation; however, the difference was more pronounced in terms of recovery temperatures. A previous report showed that the palm temperature of patients with PD is lower than that of normal controls in a normal environment,^13^ and another report showed a slower recovery of FST in patients with PD than that in healthy participants after cold stimulation.^12^ The pathomechanism of these phenomena is thought to be the hypercontraction of peripheral vessels as a result of denervation hypersensitivity of the noradrenalin receptor of peripheral vessels.^19^ However, there are some reports showing that the vasoconstriction of peripheral vessels after cold stimulation is weak in patients with MSA,^18^ with a further report showing that the vasoconstrictive ability of the skin sympathetic nerves is weak in patients with MSA.^20^ These phenomena are thought to be caused by autonomic nerve disturbances in the hypothalamus or medulla. These findings suggest that the difference in FST recovery after the 10sec-CWST between the PD and MSA groups in the present study may be due to the difference in peripheral vasoconstrictor responses derived from differences in ANS disturbance.

The third finding was that the degree of FST was negatively correlated with the degree of resting tremor. Clinically, PD has been classified into three motor subtypes: tremor-dominant (TD), postural instability of gait disturbance (PIGD), and intermittent.^21^

This difference is assumed to be related to the spread of Lewy bodies and the disturbance of various nervous systems other than the nigrostriatal system.^22^ These subtypes are not limited to differences in motor symptoms, but have also been found to be associated with a variety of non-motor symptoms. Although there have been no previous reports to our knowledge on the relationship between hand temperature and motor subtypes in PD, Gu et al.^23^ studied the relationship between autonomic symptoms using the Scale for Outcomes in Parkinson’s disease for Autonomic symptoms and the motor subtypes of PD. They reported that patients with PIGD– and indeterminate-type PD have more sweating and thermoregulatory symptoms, whereas patients with TD– and indeterminate-type PD have more cardiovascular symptoms. The regulation of body surface temperature in the hands involves the contraction and dilation of peripheral blood vessels, as well as perspiration. Perspiration is involved in the regulation of hand surface temperature and the vasoconstrictor response noted above. It is important to note that the participants in this study may have had their tremors modified by dopamine replacement therapy (DRT), and it cannot be ruled out that patients with a better response to treatment with DRT may have had a faster recovery in FST. The present study did not examine the mechanism by which differences in FST recovery occur. The involvement of vasoconstriction and sweating and the effect of reactivity to DRT are topics for future research.

This study had some limitations. The patients in our study were clinically diagnosed and no pathological diagnosis was made. However, the accuracy of the clinical diagnosis was increased by analyzing only patients whose diagnosis remained unchanged during more than 5 years of follow-up after disease onset. The number of patients with PD and MSA in this study was small, and there were no patients with typically discolored, cold hands or feet in either group. No healthy participants were included in this study. We intend to increase the number of cases in future studies and conduct comparative studies with healthy participants and patients with parkinsonian syndromes. Because this study included only Japanese patients, we do not know whether similar findings can be obtained in other ethnic groups. However, it may be possible to extract characteristics of Japanese patients with PD and MSA. In conclusion, the 10sec-CWST could be performed safely in patients with PD and MSA, and there were significant differences in the change in FST after the 10sec-CWST between these two groups. The 10sec-CWST may be an efficient test for differentiating patients with PD and MSA.

## Data Availability

The original image of the thermal imaging camera is disclosed upon request. The finger surface temperature data are presented in Supplementary Table 1.

## Acknowledgment

We thank Editage (http://www.editage.com) for English language editing.

## Author Roles

1. Conceptualization
2. Formal analysis
3. Investigation
4. Writing: A. Original draft, B. Review and editing

MT: 1, 2, 3, 4A, 4B

WH: 3

SI: 3

KA: 3

TM: 4B

AI: 3, 4B

SO: 3, 4B

## Ethical Compliance Statement

We confirm that we have read the Journal’s position on issues involved in ethical publication and affirm that this work is consistent with those guidelines. This study was approved by the Institutional Review Board of the Human Research Ethics Committee of Kanto Central Hospital (Approved number 28-5-002). Written informed consent was obtained from all participants before enrollment.

## Availability of Data

The original image of the thermal imaging camera is disclosed upon request. The FST data are presented in Supplementary Table 1.

## Legends for Figures and Supplemental Files

**Supplemental Figure 1.** A: Ipsilateral side of cold water stress test (CWST); B: Contralateral side of CWST. CWST, cold water stress test; MSA, multiple system atrophy; PD, Parkinson’s disease

