## Supplementary figures and images for "Ten-Second Cold Water Stress Test Differentiates Parkinson’s Disease From Multiple System Atrophy: A Pilot Study"

### supplemental figure 1

Supplemental Figure 1A

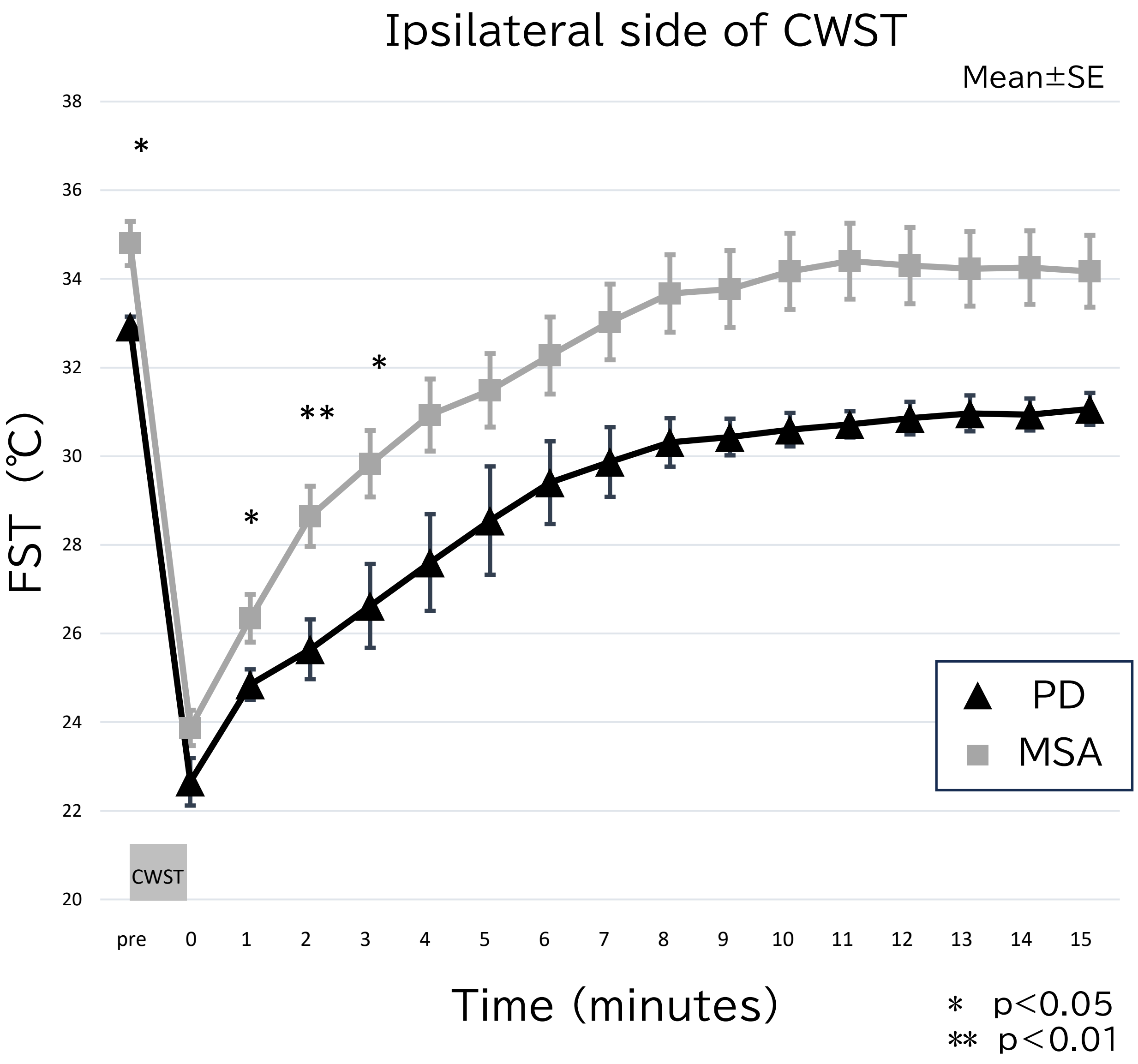


Supplemental Figure 1B

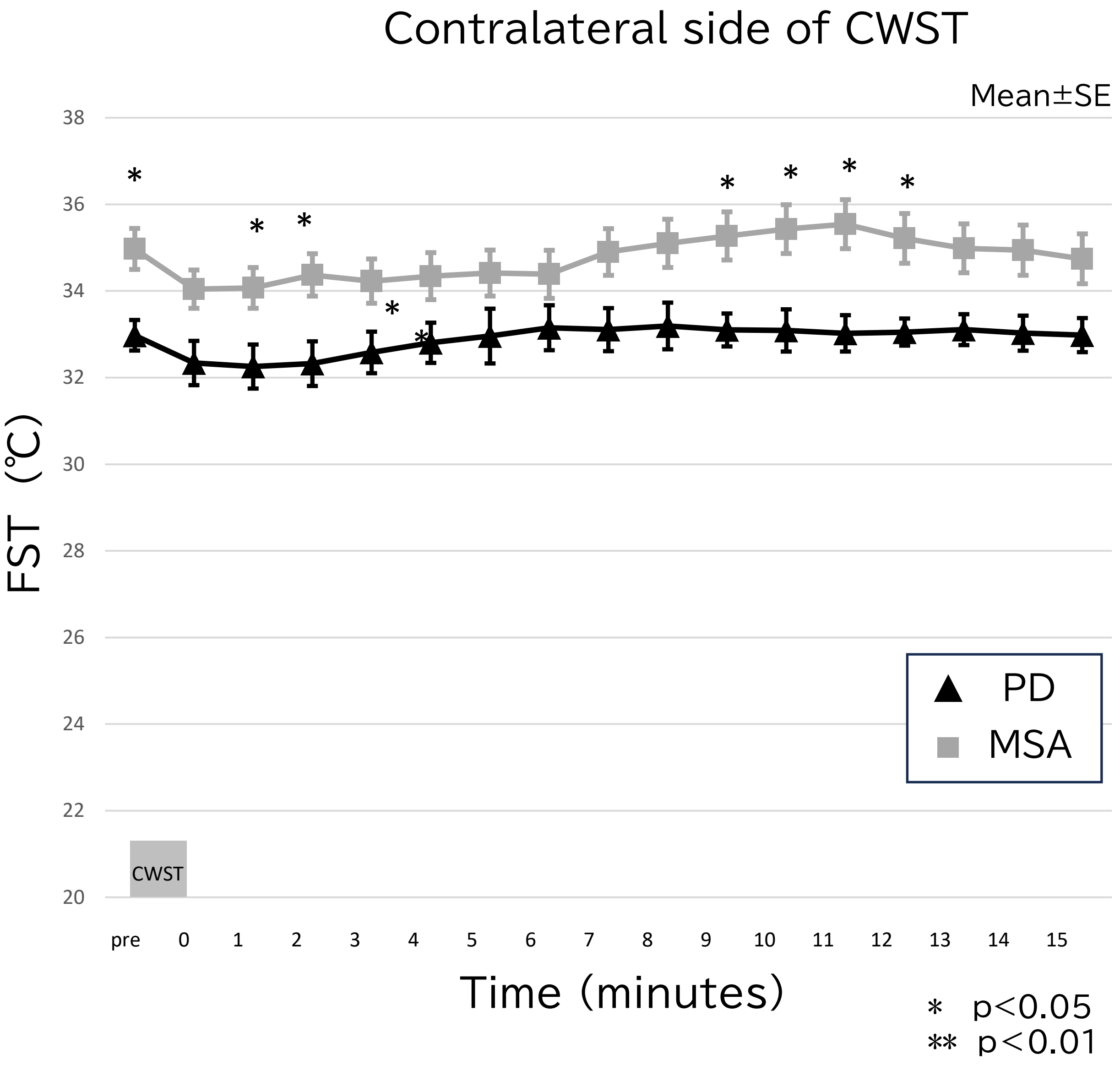
